# A Sensitive, Rapid, and Portable CasRx-based Diagnostic Assay for SARS-CoV-2

**DOI:** 10.1101/2020.10.14.20212795

**Authors:** Daniel J Brogan, Duverney Chaverra-Rodriguez, Calvin P Lin, Andrea L Smidler, Ting Yang, Lenissa M. Alcantara, Igor Antoshechkin, Junru Liu, Robyn R Raban, Pedro Belda-Ferre, Rob Knight, Elizabeth A Komives, Omar S. Akbari

## Abstract

Since its first emergence from China in late 2019, the SARS-CoV-2 virus has spread globally despite unprecedented containment efforts, resulting in a catastrophic worldwide pandemic. Successful identification and isolation of infected individuals can drastically curtail virus spread and limit outbreaks. However, during the early stages of global transmission, point-of-care diagnostics were largely unavailable and continue to remain difficult to procure, greatly inhibiting public health efforts to mitigate spread. Furthermore, the most prevalent testing kits rely on reagent- and time-intensive protocols to detect viral RNA, preventing rapid and cost-effective diagnosis. Therefore the development of an extensive toolkit for point-of-care diagnostics that is expeditiously adaptable to new emerging pathogens is of critical public health importance. Recently, a number of novel CRISPR-based diagnostics have been developed to detect COVID-19. Herein, we outline the development of a CRISPR-based nucleic acid molecular diagnostic utilizing a Cas13d ribonuclease derived from *Ruminococcus flavefaciens* (CasRx) to detect SARS-CoV-2, an approach we term SENSR (Sensitive Enzymatic Nucleic-acid Sequence Reporter). We demonstrate SENSR robustly detects SARS-CoV-2 sequences in both synthetic and patient-derived samples by lateral flow and fluorescence, thus expanding the available point-of-care diagnostics to combat current and future pandemics.

## Introduction

Following emergence from China in late 2019 ^1–3^, severe acute respiratory syndrome coronavirus 2 (SARS-CoV-2) ^1,4^ has spread to almost every country despite unprecedented control efforts ^5^. Compared to H1N1, Ebola, MERS, and SARS-CoV-1 outbreaks of recent decades, this novel coronavirus represents the first pandemic characterized by widespread global transmission coupled with significant mortality. Pre-symptomatic and asymptomatic carriers have been identified as major contributors to the prolific spread of SARS-CoV-2 ^4,6,7^ however, in many cases these patients go unidentified and unisolated, exacerbating the spread of the disease. Thus, robust identification and isolation of all infected individuals are essential for controlling disease transmission. The SARS-CoV-2 pandemic thus presents an unparalleled global public health emergency, which spurred the urgent development of molecular diagnostics and therapeutics for timely patient identification, isolation and treatment. The economic, health, and societal damage wrought by SARS-CoV-2, highlights the importance of expanding and improving on current diagnostic technologies to identify and prevent future pandemics.

Early diagnostics detected SARS-CoV-2 infection through the amplification of viral RNA (vRNA) by real-time reverse transcription polymerase chain reaction (RT-PCR) ^8–10^. These tests were time consuming ^11^, limited by reagents ^12^, required advanced equipment, and yielded significant false-negatives ^13–16^ possibly exacerbated by genetic variation within the targeted viral genomic sequences ^17^. Next generation sequencing-based diagnostics reduced false-negative rates, but still require specialized equipment and are slow (∼12 hours) ^18^. Therefore, developing alternative technologies with the potential to yield cost- and time-effective point-of-care diagnostics demands investment.

CRISPR-Cas nucleases can be easily programmed to target nucleic acids in a sequence-specific manner ^19–21^, making them prime candidates for the detection and diagnosis of viral genetic material, and forming the CRISPR-based diagnostics (CRISPRDx) pipeline ^22–25^. These systems rely on Type II Cas enzymes to physically bind target sequences ^26^, or collateral cleavage by Type V or Type VI enzymes to detect DNA ^24,25,27^ or RNA species, respectively ^22,23,28^. Since pandemic onset, an array of innovative diagnostics and prophylactics relying on these technologies have been adapted to detect or target SARS-CoV-2 with unprecedented speed ^26,29–40^, most notably represented by the DETECTR (DNA Endonuclease Targeted CRISPR Trans Reporter) ^24,25^ and SHERLOCK (Specific High-Sensitivity Enzymatic Reporter unLOCKing) ^22,23^ systems (Summarized in **Fig. S1, Table S1**).

The SHERLOCK system combines isothermal amplification of target sequences, followed by target recognition via *Leptotrichia wadei* Cas13a (LwaCas13a) and collateral cleavage of a bystander ssRNA probe to report the presence of a target ^22^. This system has undergone significant optimization since its first development in 2017. This includes improvement of *i)* sensitivity, by the inclusion of an accessory protein to amplify signal or substitution of RPA with LAMP ^23,41,42^, *ii)* specificity, by primer and guide optimization ^22,23^, *iii)* throughput, by multiplexing detection using additional enzymes (including a cocktail of LwaCas13a, PsmCas13b (*Prevotella* sp. MA2016), CcaCas13b (*Capnocytophaga canimorsus* Cc5), and AsCas12a (*Acidaminococcus* sp. BV3L6)) ^23^, and *iv)* validation as a point-of-care diagnostic by using lateral flow and ultrafast RNA extraction methods ^23,31,33,43^. Ideally, to maximize all the capabilities of SHERLOCK and expand the CRISPRDx toolkit, it is important to evaluate alternative Cas enzymes that can complement or supplement the system.

Similar to Cas ribonucleases used in other CRISPRDx systems, Cas13d enzymes such as RfxCas13d (CasRx), exclusively target RNA species that trigger subsequent collateral cleavage of bystander RNA ^44–46^. Collateral cleavage is initiated following on-target ssRNA cleavage by the HEPN domain-based endoRNase heterodimer, which activates *trans*-cleavage of nonspecific bystander RNAs ^20,44,46,47^. Furthermore, Cas13d enzymes are approximately 20% smaller than Cas13a-Cas13c effectors, and do not require a Protospacer Flanking Sequence (PFS) ^20,44,46,48^, presenting an advantage for protein production and flexible targeting. While the genetic modulatory effects of CasRx have been thoroughly characterized in *Drosophila, zebrafish*, and human cells ^44,45,49^, and its putative prophylactic properties against SARS-CoV-2 have been demonstrated ^40^, its potential as a diagnostic system has not yet been explored.

In an effort to expand the CRISPRDx toolkit, herein, we report the first use of CasRx ^44^ as a molecular diagnostic, developing a unique system we term SENSR (Sensitive Enzymatic Nucleic-acid Sequence Reporter) and demonstrating robust detection of SARS-CoV-2 viral sequences (**Fig. 1A**). Following on-target cleavage, CasRx exhibits collateral cleavage of off-target nucleic acids ^44,45^, a feature shared by Cas nucleases used in other CRISPRDx systems ^22–25^. We exploit the collateral cleavage activity of CasRx to detect SARS-CoV-2 in both synthetic templates as well as in patient-derived samples via fluorescence-based readout, and paper-based lateral flow assay. To maximize specificity, we performed an extensive bioinformatic analysis to identify novel conserved and specific viral targets to minimize false-negative and false-positive rates, respectively. We demonstrate that SENSR facilitates attomolar sensitivity in just under two hours total reaction time. The detection limit of SENSR is comparable to, though slightly less than, previously established CRISPRDx systems ^31,35,38^ and shows promise for improvement.

**Figure 1.**
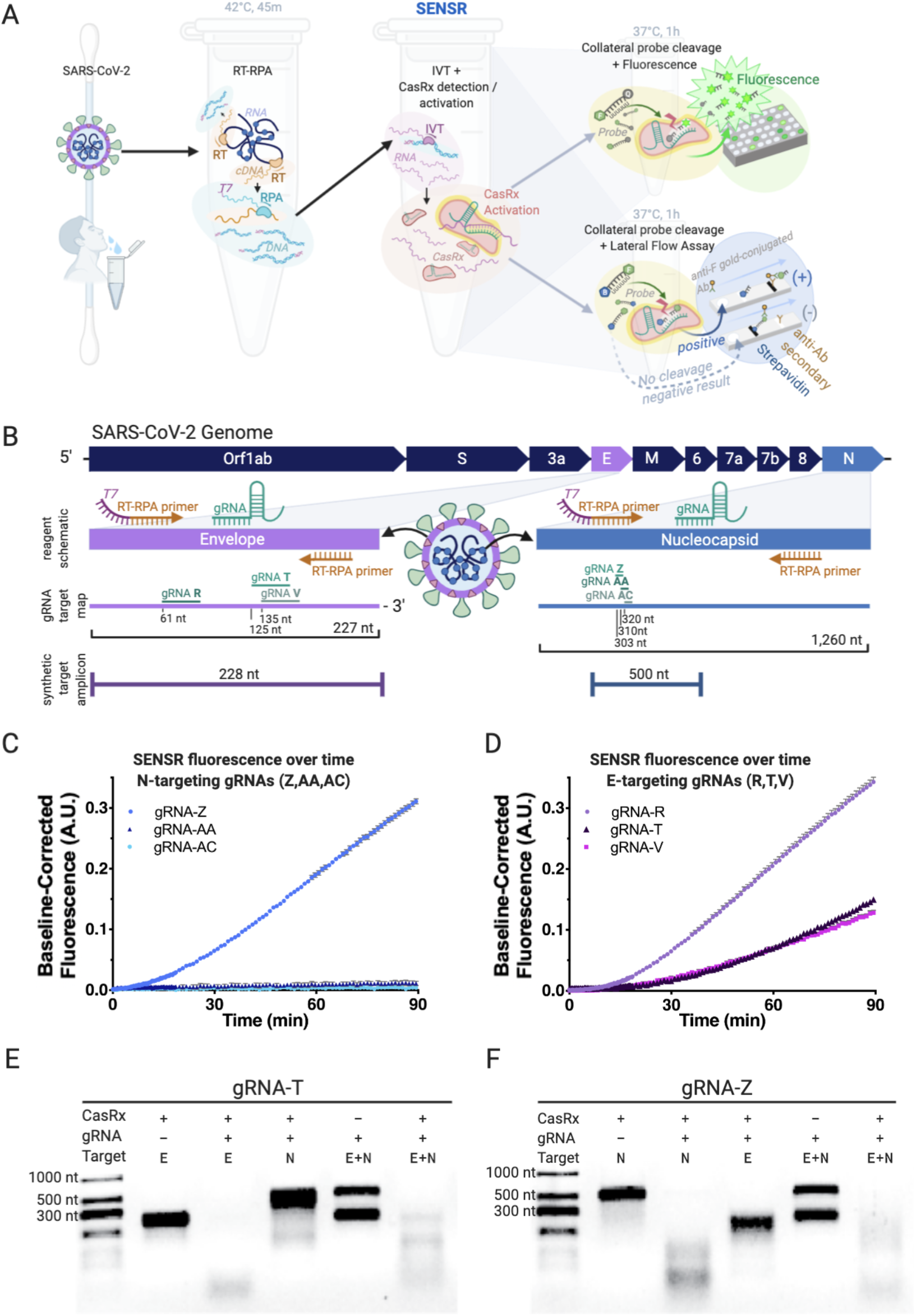
Harnessing SENSR to detect evidence of SARS-CoV-2 viral transcripts. **[A]** Overview of assay workflow. Following extraction of viral RNA, the detection protocol requires three distinct steps, the last of which differs based on desired output detection method. In the first reaction, specific target sequences within the viral RNA are reverse transcribed (RT) into cDNA and amplified by RPA at 42°C for 45m, while also adding T7 promoter sequences to the 5’ terminus (T7, purple). In the next reaction, *in vitro* transcription occurs simultaneously with CasRx collateral cleavage activation (pink) by recognition and cleavage of the target RNA sequence (purple) through the sequence-specific targeting activity of the gRNA (emerald). Addition of a probe conjugated to fluorescein and a quencher can facilitate readout by fluorescence following probe cleavage (top right). Addition of a probe conjugated to fluorescein and biotin facilitates readout by lateral flow assay (bottom right). [**B**] Schematic of the SARS-CoV-2 genome, RPA reagents, gRNA target sites, and synthetic amplicon position. The E (envelope) and N (nucleocapsid) genes are enlarged to depict the design schematic in more detail (purple and blue respectively). The schematic depicts the relative position of the RT-RPA primers and gRNAs used in the amplification step. The gRNA target map outlines all six gRNAs tested through the course of this work, with their relative positions. The E-gene encompases 227 total nt while the N-gene encompases 1,260 nt, and gRNA lengths and positions are shown to scale. Synthetic target amplicon denotes the length of the synthetic viral genome fragment used to test assay cleavage and its relative position within the gene coding sequence. **[C]** Preliminary characterization of CasRx fluorescence detection by N-targeting gRNAs (gRNA-Z, -AA, -AC) with the addition of 300 fM dsDNA template (N-gene template, 524bp) to an IVT-coupled cleavage reaction. Shown as Background Subtracted Units, A.U. over time (4 technical replicates each, Mean and SEM). gRNA-Z was selected for further downstream analysis. **[D]** Preliminary characterization of CasRx fluorescence detection by E-targeting gRNAs (gRNA-R, -T, -V) with the addition of 300 fM dsDNA template (E-gene template, 253 bp) to an IVT-coupled cleavage reaction. Plotted as Background Subtracted Units, A.U. over time (4 technical replicates each, Mean and SEM). gRNA-T was selected for further downstream analysis. **[E]** The *in vitro* cleavage properties of the E-targeting gRNA-T. In Lane 1 the absence of gRNA-T results in no cleavage of the E-gene fragment (228 nt). In Lane 2 the addition of gRNA-T results in cleavage of the E-gene template resulting in loss of the distinct 228 nt band and appearance of lower molecular weight cleavage species. In Lane 3, the off-target N-gene template (500 nt) remains uncleaved in the presence of CasRx and the E-targeting gRNA-T. In Lane 4, the absence of CasRx results in no cleavage of either E or N-gene templates. In Lane 5, the on-target cleavage of the E-gene template results in additional collateral cleavage of the N-gene template, resulting in the loss of both bands from the gel and the accumulation of lower molecular weight cleavage products. **[F]** The *in vitro* cleavage properties of the N-targeting gRNA-Z. In Lane 1, the absence of gRNA-Z results in no cleavage of the N-gene fragment (500 nt). In Lane 2, the addition of gRNA-Z results in cleavage of the N-gene template resulting in loss of the distinct 500 nt band and appearance of lower molecular weight cleavage species. In Lane 3, the off-target E-gene template (228 nt) remains uncleaved in the presence of CasRx and the N-targeting gRNA-Z. In Lane 4, the absence of CasRx results in no cleavage of either E or N-gene templates. In Lane 5, on-target cleavage of the N-gene template results in additional collateral cleavage of the E-gene template, resulting in the loss of both bands from the gel and the accumulation of lower molecular weight cleavage products.

## Results

### Development of the SENSR protocol

Derived from protocols originally developed for (**Fig. S1, Table S1**) SHERLOCK ^22,23,31,48^, we designed a two-step protocol for the detection of nucleic acids using recombinant CasRx purified after expression in *E. coli* (**Fig. S2**). The protocol requires an initial 45-minute isothermal preamplification reaction combining reverse transcriptase and recombinase polymerase amplification (RT-RPA) ^50^ producing a short dsDNA amplicon encompassing the CasRx target site and containing a T7 promoter sequence. This is followed by an *in vitro* transcription (IVT)-coupled cleavage reaction which converts the dsDNA amplicons into ssRNA, recognizable by CasRx for cleavage, resulting in collateral cleavage of a bystander ssRNA probe. Collateral cleavage of the probe is analyzed by either fluorescence or lateral flow readouts, thus indicating the initial presence or absence of SARS-CoV-2 genomic sequences (**Fig. 1A, Fig S3**).

### Target selection and reagent validation

To ensure high analytical specificity, we identified conserved gRNA target sites using a bioinformatic pipeline to identify 30 nt target sequences conserved among the first 433 published SARS-CoV-2 genomes available at Genbank on April 7^th^, 2020, and without homology to other coronaviruses (ViPR, Virus Pathogen Resource, n = 3,164). This yielded a panel of gRNA target sites (n=8846) less likely to result in false positives or negatives due to sequence constraints (**Fig. S4, Table S2, S3)**. Because RT-qPCR assays recommended by the WHO and CDC target the envelope (E) and nucleocapsid (N) genes within the SARS-CoV-2 genome, we selected these genes as the targets of SENSR for system validation (**Fig. 1B**) ^8,51^. The bioinformatic analysis revealed multiple specific sequences within the N-gene (n = 150), to which we designed three gRNAs (gRNA-Z, -AA, -AC) (**Table S2, Fig. S5A**). However, the stringent bioinformatic search criteria did not identify targets specific to SARS-CoV-2 within the E-gene due to high sequence homology with the SARS-CoV-1 E-gene. We, therefore, relaxed the criteria to include target sites sharing homology with distantly related SARS-CoV-1 which is presently absent from the general population. From this, we selected three E-targeting guides (gRNA-R, -T, -V), with two displaying complete (gRNA-T (433/433), gRNA-V (433/433)) and one with nearly complete (gRNA-R (430/433)) conservation among the available SARS-CoV-2 genomes (**Table S4, Fig. S5A)**. To validate these gRNAs, we generated synthetic SARS-CoV-2 targets encompassing the E- and N-genes **(Table S5)**, and tested for *in vitro* cleavage with purified CasRx protein (**Fig. S2**). Initial characterization of on-target cleavage properties revealed significant degradation of target transcripts for all guides tested (**Fig. S5B, C**) motivating further assessment of all candidates. To determine the most effective gRNAs for use in SENSR, we monitored fluorescence accumulation over time in an IVT-coupled cleavage reaction for each gRNA. All gRNAs induced robust fluorescence within minutes, with the exception of gRNA-AA and -AC which produced no signal (**Fig. 1C, D**). gRNA-T (E-targeting) and gRNA-Z (N-targeting) were selected for downstream analysis due to their robust cleavage activity as well as sequence conservation among all publicly available SARS-CoV-2 patient sample isolates (**Fig. 1B-D, Table S2, S3**).

### Fluorescence-based detection of SARS-CoV-2

We and others have recently demonstrated that on-target cleavage activates a secondary collateral cleavage property of CasRx ^44,45^. We initially evaluated the *in vitro* collateral cleavage activity of CasRx with gRNA-T and gRNA-Z through gel electrophoresis. By incubating CasRx, gRNA-T or gRNA-Z, and varying the addition of synthetic templates, we found CasRx collateral cleavage was only activated when the synthetic template added complemented the gRNA target sequence (**Fig. 1E, F**). We sought to exploit this tandem, collateral cleavage activity to cleave a bystander fluorescent probe in *trans*, facilitating detection of SARS-CoV-2 by fluorimetry **(Fig. 1A, Fig. S3)**. The quenched fluorescent reporter within RNaseAlert v2 has been used to report collateral cleavage of *Leptotrichia wadeii* (LwaCas13a), *Capnocytophaga canimorsus Cc5* (CcaCas13b), and PsmCas13b (*Prevotella* sp. P5-125) in the SHERLOCK systems ^22,23,48^, but our preliminary analysis of RNaseAlert v2 failed to yield fluorescence with CasRx in the presence of a synthetic target (Student’s t-test gRNA-T: p=0.4294, gRNA-Z: p=0.1510) (**Fig. S6A**). Prior work suggested CasRx preferentially cleaves targets containing poly-U stretches ^44^, with complementary work demonstrating collateral cleavage of targets rich in both poly-A and poly-U stretches ^46^. To develop a probe cleavable by CasRx, we therefore generated two custom 6 nucleotide ssRNA probes, poly-A and poly-U, each conjugated to a 5’fluorescent molecule (6-FAM) and a 3’fluorescence quencher (FQ), whose separation following cleavage results in detectable fluorescence accumulation (**Fig. 1A**). Following incubation of CasRx, with either gRNA-T or -Z, in the presence or absence of their respective synthetic targets (E- or N-gene), the poly-U probe yielded significant fluorescence for 10000 copies/*μ*L compared to the no-template control (NTC) (Student’s t-test gRNA-T, gRNA-Z: p<0.0001), while the poly-A probe produced no detectable fluorescence signal (Student’s t-test gRNA-T: p=0.5953, gRNA-Z: p=0.7935), suggesting a preference for poly-U stretches by CasRx, and motivating its use in the remainder of experiments (**Fig. S6A, Table S5**).

Following probe selection, we moved to evaluate the collateral cleavage activity in the context of fluorescence. We set to determine if the gRNA incubated with the respective target sequence dictates the increase in fluorescence signal over time. To do so, we incubated CasRx, gRNA-T or gRNA-Z, the modified poly-U probe, and varied the addition of synthetic templates, while fluorescence data were acquired on a plate reader. We observed fluorescence signal only accumulated, and thus collateral cleavage activated, when the synthetic template added to the reaction complemented the gRNA target sequence (**Fig. 2A, B**). After optimizing preamplification reaction input volume using 100 copies/*μ*L of synthetic RNA, and determining 50% preamplification reaction volume input to be optimal (**Fig. S6B**), we then determined the LOD by fluorescence for gRNA-T and gRNA-Z titrated at log scale from 10,000 to 0 copies/*μ*L. We determined the LOD to be 100 copies/*μ*L (**Fig. 2C, D**), indicating SENSR exhibits attomolar sensitivity comparable to other CRISPRDx systems ^22,24^. These results demonstrate CasRx can robustly detect and report the presence of synthetic SARS-CoV-2 RNA via fluorescence readout.

**Figure 2.**
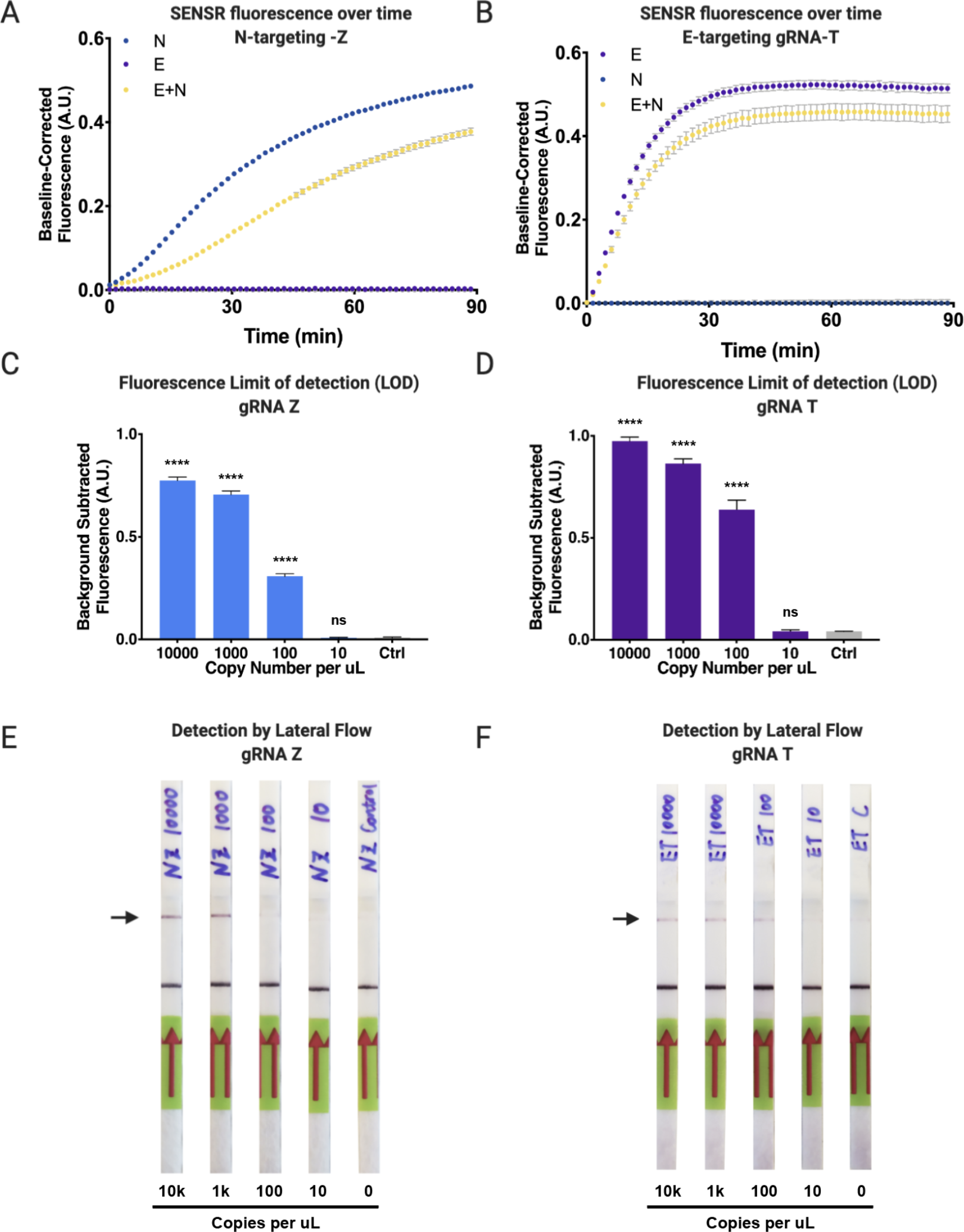
SARS-CoV-2 detection by SENSR via Fluorescence and Lateral Flow assay. **[A]** Cleavage properties of E-targeting gRNA-T in fluorescence context. Fluorescence detection of E-gene in an IVT-coupled coupled context. gRNA-T incubated in three different contexts: with E-gene template, with N-gene template, or with both E- and N-gene template. Accumulation of fluorescence occurs when gRNA-T is incubated with the E-gene target template leading to cleavage of the ssRNA probe. Collateral cleavage is not observed when gRNA-T is incubated with the non-target N-gene template. **[B]** Cleavage properties of N-targeting gRNA-Z in fluorescence context. Fluorescence detection of N-gene in an IVT-coupled coupled context. gRNA-Z incubated in three different contexts: with N-gene template, with E-gene template, or with both N- and E-gene template. Accumulation of fluorescence occurs when gRNA-Z is incubated with the N-gene target template leading to cleavage of the ssRNA probe. Collateral cleavage is not observed when gRNA-Z is incubated with the non-target E-gene template. **[C]** CasRx nucleic acid limit of detection (LOD) for gRNA-Z detection via fluorescence following cleavage of N-gene target. Total copy number of 10000, 1000, 100, 10, or 0 of viral RNA template input into initial RT-RPA reaction, followed by CasRx detection by fluorescence. Performing a one-way ANOVA followed by a Dunnett’s test call copy numbers to the NTC significance was found for 10000, 1000, and 100 copies (p<0.0001) and no significance was found for 10 copies (p=0.9999). Results shown as background-subtracted fluorescence (A.U.) following 90 minute fluorescent readout (4 technical replicates each, Mean and SD). **[D]** CasRx nucleic acid LOD for gRNA-T detection via fluorescence following cleavage of E-gene target. Total copy number of 10000, 1000, 100, 10, or 0 of viral RNA template input into initial RT-RPA reaction, followed by CasRx detection by fluorescence. Performing a one-way ANOVA followed by a Dunnett’s test call copy numbers to the NTC significance was found for 10000, 1000, and 100 copies (p<0.0001) and no significance was found for 10 copies (p>0.9999). Results shown as background-subtracted fluorescence (A.U.) following 90 minute fluorescent readout (4 technical replicates each, Mean and SD). **[E]** LOD of the gRNA-Z recognition of N-gene target by lateral flow assay. CasRx detection reaction incubated for 1h prior to lateral flow assay. **[F]** LOD of the gRNA-T recognition of E-gene target by lateral flow assay. CasRx detection reaction incubated for 1h prior to lateral flow assay.

### Lateral flow assay development

Collateral cleavage by CasRx can additionally be exploited to detect synthetic SARS-CoV-2 RNA by lateral flow assay, facilitating detection by simple paper test strip and eliminating the need for expensive laboratory equipment **(Fig. 1A, Fig. S3)**. Similar to assays developed by others ^23,48^, we developed a 6 nucleotide ssRNA probe conjugated with 5’6-FAM and 3’biotin (Bio) compatible with Millenia HybriDetect lateral flow strips (**Table S5**). In brief, collateral cleavage results in separation of 6-FAM from biotin, detectable following capillary action down a paper dipstick imprinted with streptavidin or anti-FAM secondary antibodies at distant ends (**Fig. 1A**). The absence or presence of the upper band therefore indicates a negative or positive result, respectively (Summarized in **Fig. 1A, Fig. S3**). Using this protocol, we demonstrated that SENSR can be adaptable to readout by lateral flow, and determined the LOD of synthetic SARS-CoV-2 RNA to be as low as ∼100 copies/*μ*L, however with variability between guides (**Fig. 2E, F**). These results confirm that SENSR, like other CRISPRDx systems, can be adapted for read-out by lateral flow, indicating the potential for future application within point-of-care rapid diagnostic tests.

### Specificity of SENSR against known possible off-targets

Diagnostic assays require stringent specificity parameters to limit false-negatives/positives. Because many Cas effectors tolerate some degree of mismatch ^52–54^, unintended false-positives can occur as a result of cleaving closely related off-target sequences. In a health-care setting, SENSR is unlikely to be exposed to randomly generated high-identity sequences, and will more likely encounter closely related natural homologs. Therefore, we identified the four highest-identity natural homologous sequences to the gRNA-T and gRNA-Z target sites via BLAST. In each case, SARS-CoV-1 variants, Bat coronaviruses, and Pangolin coronaviruses were identified as the most closely related potential off-targets (OT), containing 2 or 3 mismatches, with gRNA-Z also targeting an additional unknown marine virus and a porcine genome sequence with 7 mismatches (**Fig. S7A, B, Table S6**). We synthesized these sequences as DNA templates containing a T7 promoter and assayed for off-target cleavage tolerance of gRNA-T and gRNA-Z in an IVT-coupled cleavage reaction analyzed via fluorescence detection. We found that gRNA-T was sufficiently specific to exclusively recognize the synthetic E-gene fragment (ANOVA, p<0.001), precluding recognition of any off-targets, which showed no significant differences in fluorescent signal with the NTC (ANOVA, Dunnett’s test OT1: p=0.9998, OT2: p=0.5242, OT3: p=0.6633, OT4: p=0.8475) (**Fig. S7C**). gRNA-Z demonstrated detection of the N-gene synthetic template and the most similar pangolin coronavirus off-target templates compared to the NTC (N, OT1, OT2: p<0.0001), thus presenting some degree of mismatch tolerance for gRNA-Z. Fluorescence signal for OT3 and OT4 did not differ from the NTC (ANOVA, Dunnett’s test OT3: p=0.8877, OT4: p>0.9999) (**Fig. S7D**).

### CasRx-based detection of SARS-CoV-2 from patient isolates

We next sought to determine the capability of SENSR to detect SARS-CoV-2 from infected patient samples and directly compared these results to RT-qPCR-validated diagnostics. RT-qPCR analysis of patient samples was performed by targeting the N-, S-, and Orf1ab-genes (**Table S7**), and accordingly, we selected gRNA-Z to directly compare SENSR fluorescence detection to N-gene RT-qPCR Ct values. We performed fluorescence detection analysis on 42 RT-qPCR validated positive (n = 21) and negative (n = 21) patient samples. By fluorescence, SENSR yielded no false-positives among negative patient samples demonstrating 100% specificity (0/21), and obtained a conservative 57% concordance with confirmed positive samples (12/21) when the threshold for detection is set at S/N > 2 (**Fig. 3A, B, Table S7**). For RT-qPCR analysis of viral infections, a lower Ct value indicates a higher viral load within an isolated sample. SENSR demonstrated robust detection of infected patient samples with Ct values ≤ 20, detecting SARS-CoV-2 vRNA in (8/9) such samples (Sample ID: 1-9) (**Fig. 3A, Table S7**). SENSR could detect SARS-CoV-2 vRNA up to a maximum Ct value ≤ 28 with a moderate 25% (4/16) false-negative rate (**Fig. 3A, B, Table S7**). Next, we confirmed detection of SARS-CoV-2 can be achieved by lateral flow analysis. We performed lateral flow analysis on the 12 positive samples resulting in S/N > 2, and correlating to a Ct ≤ 28, to determine if SENSR can operate as a point-of-care diagnostic. Using lateral flow, we observed 92% (11/12) concordance with SENSR fluorescence analysis (**Fig. 3C**). These findings confirm SENSR can be successfully adapted for the rapid detection of SARS-CoV-2 from patient samples, however, further optimization will be required to improve sensitivity and consistency.

**Figure 3.**
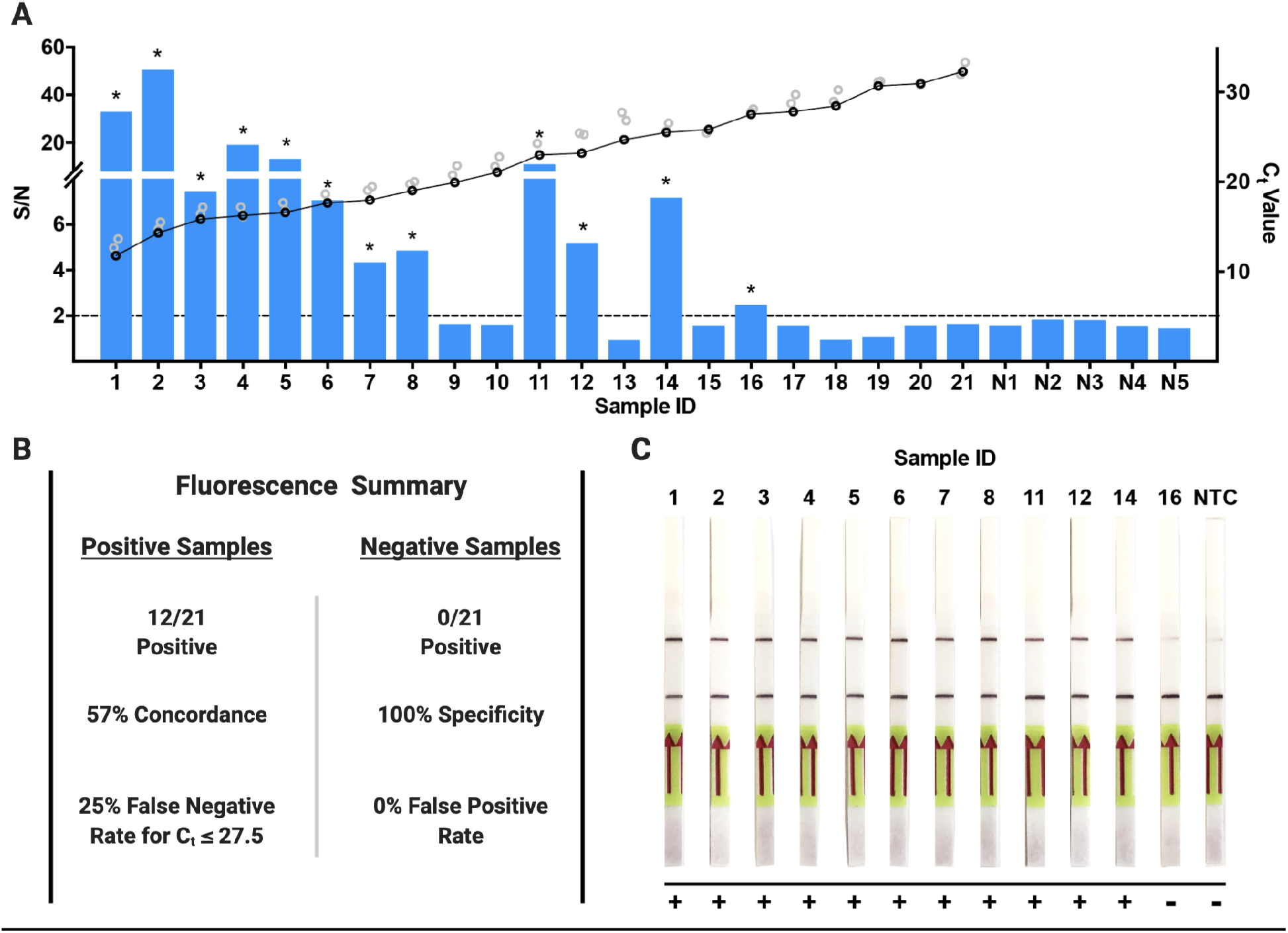
SENSR detection of positive SARS-CoV-2 validated patient samples. **[A]** SENSR fluorescence analysis of RT-qPCR validated patient samples using gRNA-Z for detection. Samples 1-21 are patient samples positive for SARS-CoV-2 listed in order of ascending RT-qPCR Ct values for the N-gene where low Ct value is equal to a high viral load. The black and gray rings represent the Ct values for N, S, and Orf1ab genes, where black represents the N-gene Ct values and gray represents the S and Orf1ab Ct values. Samples N1-N5 are the five samples negative for SARS-CoV-2 with the highest recorded signal-to-noise ratios (S/N) of all negative samples analyzed with the highest S/N = 1.8. The dashed line represents the S/N = 2 threshold used to determine a positive detection of SARS-CoV-2. The asterisks indicate a positive detection of SARS-CoV-2 in the patient sample. **[B]** Summary of fluorescence detection results for the RT-qPCR validated positive and negative patient samples. **[C]** Lateral flow based detection of the 12 samples from **[A]** that resulted in a positive detection of SARS-CoV-2 (S/N > 2). The top band represents the test band and the bottom the control band. An increase in saturation of the top band indicates a positive detection of SARS-CoV-2 in the sample. A positive result was also determined by comparing to the NTC, which served as a baseline for a negative result.

## Discussion

With an increasingly interconnected world and expanding global population, future pandemics are inevitable. The COVID-19 pandemic spread prolifically in the early months of 2020, with containment elusive in part due to the scarcity of point-of-care diagnostics. The seemingly infinite adaptability of CRISPR has, or promises to, accelerate the development of everything from life-saving gene therapies ^55–57^ and pig-to-human organ donations ^58^; to disease-eradicating gene drives ^59–61^ and possibly the re-animation of the Woolly Mammoth ^62,63^ - with CRISPR-based diagnostics (CRISPRDx) being no exception. Though still nascent, CRISPRDx, like other CRISPR technologies, has proven fast to develop, highly flexible, capable of multiplexing, making it the ideal toolkit from which to develop expeditious future point-of-care diagnostics. The CRISPRDx technologies developed prior to the COVID-19 pandemic, such as SHERLOCK and DETECTR, may have helped halt disease transmission had they been deployed earlier and implemented more widely. Therefore, it is important to prepare now, well in advance of the next pandemic, by perfecting and expanding the CRISPRDx toolkit to the bounds of its capabilities.

Complementing the rapidly expanding CRISPRDx toolkit (**Fig. S1, Table S1**), here we outline the use of RfxCas13d (CasRx) to diagnose SARS-CoV-2 using both synthetic targets and patient derived samples. SENSR amplifies nucleic acid sequences through an isothermal alternative to PCR then detects the target sequences by exploiting the native collateral cleavage activity of CasRx (**Fig. 1A**), providing proof-of-principle that Cas13d can be adapted as a point-of-care diagnostic. We lend further evidence that CasRx cleavage results in activation of an off-target collateral cleavage property (**Fig. 1 E, F**) ^44,64^, with a preference for poly-U stretches (**Fig. S6A**). This feature can be harnessed to detect viral sequences in a single reaction or possibly even in a multiplexed reaction combined with other Cas enzymes lacking a poly-U preference ^23,48^. We identify highly conserved and specific targets to SARS-CoV-2 to prevent false-negatives (**Fig. S4, Table S2, S3**), and demonstrate detection with attomolar sensitivity by both fluorescence and lateral flow readouts comparable to other previously developed systems (**Fig. 2C-F**) ^31,38,39,65^. We further demonstrate that SENSR has the potential to be so specific to SARS-CoV-2, and further prove it can be adapted to detect vRNA in infected patient isolates (**Fig. 3A-C**).

SENSR provides a robust proof-of-principle of viral detection by CasRx, and as such, it should be considered a candidate diagnostic system; however, it requires optimization in advance of deployment. Optimizing SENSR diagnostics can be pursued through a number of avenues. While some groups have improved specificity by selectively generating synthetic mismatches in guide sequences ^22^, the gRNAs tested herein have moderate target specificity (**Fig. S7**). RT-RPA also exhibits nonspecific amplification ^66–68^, and therefore, alternative isothermal amplification technologies, such as RT-LAMP ^41,69^, could be implemented to improve specificity. Although RT-LAMP is likely to improve specificity as well as sensitivity, the requirement of two separate reactions remains problematic, increasing the likelihood of contamination due to sample transfers ^70^. RT-LAMP and CasRx operate optimally at divergent temperatures (60-65°C and 37°C, respectively), which may be incompatible within a one-step molecular diagnostic, though advances have been made in this sphere ^33,71,72^. Thus alterations to existing isothermal amplification technologies or discovery of novel thermostable Cas nucleases with collateral cleavage activity could facilitate a more highly-sensitive Cas-based molecular diagnostic that operates in one reaction at a single temperature.

Beyond amplification, improvement to gRNA design criteria could drastically improve gRNA selection for diagnosis and consequently the response time to future disease outbreaks. Currently, there remains no robust study attempting to characterize the *in vitro* collateral cleavage activity for varying Cas13 gRNA sequences, thus limiting efficient gRNA design and target selection for Cas13-based diagnostics. In this study, we observed significant variation in gRNA collateral cleavage activity, including two gRNAs (gRNA-AA and gRNA-AC) incapable of producing fluorescence signal (**Fig. 2A**) and kinetic variation exemplified by temporal differences in fluorescence signal accumulation for gRNA-T and gRNA-Z (**Fig. 1E, F**). We also observed mild off-target activity for gRNA-Z (**Fig. S7D**) and found variation in reporter saturation for lateral flow between gRNAs (**Fig. 2E, F**), which has been found in other systems ^31,38^. Understanding gRNA-specific differences of Cas13 collateral cleavage will allow the development of functional gRNA target sequence libraries for use in future pandemics. Furthermore, robust exploration for CasRx gRNA truncations and permutations should be undertaken to generate gRNAs intolerant of target site polymorphisms, or to even distinguish between strains ^22^, thus improving the specificity of SENSR. Optimizing workflow, deployment, and distribution, while taking steps to reduce the risk of contamination, is imperative to develop CasRx-based diagnostics to their full potential. Although optimization is required, we demonstrate that detection with CasRx is a promising advancement for detecting viral infections, and could be improved to become a powerful molecular diagnostic with numerous applications.

CasRx-based diagnostic systems may present a worthy advancement for CRISPRDx due to the fundamental characteristics of the Cas13d family. Like LwaCas13a, Cas13d is more flexible than most other Cas enzymes because it lacks a protospacer flanking sequence (PFS) requirement ^28,44,46^, permitting targeting of any sequence without constraint. In addition, some native Cas13d systems include a WYL1-domain-containing accessory protein, which has been demonstrated to increase the on-target and collateral cleavage efficiency of the Cas13d effectors ^46,73^, suggesting potential for future implementation. Furthermore, because they target RNA, next-generation Cas13-based systems may be capable of direct recognition of RNA, possibly at the single molecule level, without need for a prior reverse transcription (RT) and/or amplification step. This property could enable direct detection of many emerging viral threats including, but not limited to; bunyaviruses ^74^, zoonotic viruses such as Ebola, hanta, and Lassa ^75^; arboviruses such as dengue, chikungunya, and Zika ^23,76,77^, and other coronaviruses such as MERS, SARS-CoV-1, as well as those yet undiscovered ^78,79^. CasRx-based diagnostics systems could detect endemic pathogens capable of zoonotic transmission through livestock and wild animals such as influenza or other coronaviruses ^78,80,81^ which may have been able to prevent past pandemics ^82^, and avert mass herd culling resulting in billions of dollars of losses ^83,84^. Beyond detection in patients and livestock, SENSR could be adapted to detect pathogens in insect disease vectors as well as infected individuals ^85^, facilitating rapid one-pot field detection of mosquito-borne pathogens in areas lacking laboratory infrastructure ^86^. However, SENSR is not limited to detection of RNA species, and could also be used to detect pathogen DNA (**Fig. S8**). By including an RNA polymerization step, this same technology could be harnessed to track evidence of insecticide resistance alleles ^87^, released transgenic cargoes ^59,60^, or the presence of *Borrelia burgdorferi* in a tick plucked from a hiker’s leg.

Pushing the boundaries of viral sequence recognition with CRISPR-Cas nucleases is not only of interest for genetic engineering and diagnostics, but also for therapeutics as well. The adaptability of CasRx RNA-targeting has recently been demonstrated to be a potentially powerful anti-COVID therapeutic ^40^ as well as for other viruses ^88^. Together with acute diagnostics, these technologies could promise a new mode of response to future viral outbreaks via a ‘plug-n-chug’model, in which complementary diagnostics and therapeutics could be systematically rolled out almost immediately after completion of a viral genome sequence. Similar to LwaCas13a, CasRx could also be adapted to massively multiplexed arrays to facilitate identification of viral pathogens on a large scale ^37^. Establishing these tools and frameworks now, could expedite response times and help prevent future outbreaks, avoiding the economic and health consequences which have resulted from poor preparedness to the current pandemic.

## Data Availability

All data is available in the manuscript

## Author Contributions

O.S.A. conceptualized the study. D.J.B., D.C.R., C.P.L., J.L., L.M.A., T.Y., I.A., P.B.F, performed, designed and analyzed various molecular, bioinformatic, and protein biochemistry. A.L.S, R.R., R.K., E.A.K. contributed to analyzing and designing experiments. All authors contributed to writing, analyzing the data, and approving the final manuscript.

## Data and Materials Availability

Bioinformatic scripts can be found in **File S1**. A limited quantity of CasRx enzyme will be made available upon request.

## Materials and Methods

### CasRx subcloning, protein expression and purification

To produce an expression plasmid for CasRx protein production, we cloned the human codon optimized CasRx coding sequence into the expression vector, pET-His6-MBP-TEV-yORF (Series 1-M) (purchased from QB3 MacroLab, Berkeley) using the Gibson assembly method (Gibson et al., 2009). In brief, the CasRx coding sequence was PCR amplified from plasmid OA-1050E (Addgene plasmid # 132416^64^) using primers 1136I.C1 and 1136I.C2 (**Table S5**). The fragment was purified and subcloned into the restriction enzyme cutting site EcoRI, downstream of the His-MBP recombinant protein in pET-His6-MBP-TEV-yORF, generating the final pET-6xHis-MBP-TEV-CasRx (OA-1136J; Addgene plasmid # 153023) plasmid.

Protein expression, culture, cell lysis, affinity and further downstream protein purification were performed as previously described (**Fig. S2**) ^44^. In brief, to facilitate protein expression in liquid culture, pET-His6-MBP-TEV-CasRx was transformed into Rosetta2 (DE3) pLysS cells (Novagen, 71403). Starter cultures in LB were supplemented with kanamycin and chloramphenicol and incubated at 37° C overnight. 20 mL of starter culture were used to inoculate 1L of TB media supplemented with the same antibiotics. Cultures were allowed to grow until OD600 ∼0.5, cooled on ice, induced with 0.2 mM IPTG, and then grown for 20 hours at 18° C. Cells were then pelleted, freeze-thawed, resuspended, lysed via sonication and clarified by centrifugation. Protein purification was performed by gravity Ni-NTA affinity chromatography (Thermo Scientific™HisPur™Ni-NTA Resin) followed by removal of the 6xHis-MBP tag by TEV protease concurrently with overnight dialysis. Further purification was achieved by cation exchange using a 5-mL HiTrap SP HP using a gradient of 125 mM to 2M NaCl in 50 mM Tris-HCl, 7.5% v/v glycerol, 1 mM DTT. The protein was finally purified by gel filtration chromatography in 50 mM Tris-HCl, 600 mM NaCl, 10% glycerol, 2 mM dithiothreitol on a Superdex^®^ 200 16/600 column. Fractions were pooled, concentrated to ∼ 2 mg/mL and stored at -80 C in the same buffer.

### Production of target SARS-CoV-2 RNA and gRNAs

To detect viral genomic sequences, we designed two synthetic dsDNA gene fragments containing a T7 promoter sequence upstream of gene segments corresponding to the SARS-CoV-2 envelope (E) and nucleocapsid (N) protein coding regions (GeneBank Accession # MN908947). The 253bp SARS-CoV2 E-gene segment was ordered and synthesized as a custom gBlock® from Integrated DNA Technologies (IDT) and amplified using primers 1136Q-F and 1136Q-R (**Table S5**). A 500bp SARS-CoV2 N-gene segment was amplified from a plasmid 1136Y (Catalog # 10006625) ^65^ using primers 1136X-F and 1136X-R (**Table S5**). These two SARS-CoV-2 gene targets were amplified using PCR and then purified using the MinElute PCR Purification Kit (QIAGEN #28004). We also designed eight synthetic dsDNA templates containing nucleotide variations from the native SARS-CoV-2 E- and N-gene (4 synthetic targets each gene) that were used for gRNA off-target analysis and ordered as a gBlock® from IDT. Primers 1136-OFF-F and 1136-OFF-R1∼1136-OFF-R5 were used to amplify these sequences (**Table S5**). The synthetic targets were chosen based on sequence homology identified using NCBI BLAST searches against gRNA-T and gRNA-Z. 40nt regions flanking the mismatch target sequences were included in the 5’and 3’ends of the 30 nt stretch in order to allow amplification analysis via RT-RPA.

We designed gRNAs targeting the synthetic vRNA gene segments using criteria previously outlined (**Fig. S5**) ^64^ and generated these following a previously described templateless PCR protocol ^89^. The primers used to amplify these gRNAs, as well as their final sequence are outlined in **Table S5**. We synthesized both the synthetic vRNA and gRNAs through *in vitro* transcription (IVT) using MEGAscript™ T7 Transcription Kit (Invitrogen™ #AM1334), followed by DNaseI digestion and purification using the MEGAclear™ Transcription Clean-Up Kit (Invitrogen™ #AM1908).

### *In vitro* gRNA cleavage assays

To test the *in vitro* cleavage efficiency of gRNAs, we performed preliminary *in vitro* cleavage assays to test on-target cleavage, off-target cleavage, and collateral-cleavage properties. On-target cleavage assays were prepared with RNA templates for E-gene (1000 ng) or N-gene (1500 ng), followed by addition of CasRx (112 ng) and 10 ng of each gRNA in a 2:1 molar ratio. Reactions were prepared in 20 mM HEPES pH 7.2 and 9mM MgCl_2_, incubated at 37°C for one hour, denatured at 85°C for 10 min in 2X RNA loading dye (New England Biolabs, #B0363) and loaded on 2% 1X TBE agarose gel stained with SYBR™ gold nucleic acid staining (Invitrogen #S11494). Off-target cleavage assays were assembled similarly with the non-targeting synthetic vRNA template. Collateral-cleavage assays were prepared with both synthetic vRNA templates simultaneously and same quantities of gRNA and CasRx described above.

### Bioinformatics of SARS-CoV-2 SENSR target sites

433 SARS-CoV-2 genomes were downloaded from NCBI Virus (https://www.ncbi.nlm.nih.gov/labs/virus/vssi/#/virus?SeqType_s=Nucleotide&VirusLineage_ss=SARS-CoV-2,%20taxid:2697049) and 3,164 non-SARS-CoV-2 Coronavirinae genomes were downloaded from Virus Pathogen Resource (https://www.viprbrc.org/brc/home.spg?decorator=corona_ncov) on April 7, 2020. To assess the specificity of our probes, all possible 30 nt sequences were extracted from the two genome sets using a Perl script (**File S1**) generating 52,712 and 8,338,305 unique fragments from SARS-CoV-2 and non-SARS-CoV-2 genomes, respectively. The probes designed to target E and N genes based on Corman et al. 2020 (https://www.eurosurveillance.org/content/10.2807/1560-7917.ES.2020.25.3.2000045) were cross-referenced against the extracted sequences to identify numbers of targeted genomes in each set. Four of six probes perfectly matched sequences in all 433 SARS-CoV-2 genomes. Two others, 1136R-E-Protein-gRNA1 and 1136S-N-Protein-gRNA1, matched 430 and 426 SARS-CoV-2 genomes, respectively. Of 3,164 non-SARS-CoV-2 viruses, the probes matched between 1 and 10 genomes, mostly from bat hosts (Summarized in **Table S2**). To identify a comprehensive set of possible targets that are specific to SARS-CoV-2 genomes, 16,645 30 nt sequences that perfectly matched all 433 SARS-CoV-2 genomes were filtered to remove the ones that were also found in any of the 3,164 non-SARS-CoV-2 genomes to produce a set of 8,846 SARS-CoV-2-specific sequences (**Table S4**). To check for possible cross-reactivity with human transcripts, the probes were mapped to the human transcriptome (GRCh38, ENSEMBL release 99, ftp://ftp.ensembl.org/pub/release-99/fasta/homo_sapiens/) comprising both coding and non-coding RNAs using bowtie 1.2.3 allowing up to two mismatches (-v 2). None of the 8,846 sequences mapped to the human transcriptome to confirm their specificity to SARS-CoV-2. To visualize the distribution of the specific targets along the SARS-CoV-2 genome, probe density was calculated using a sliding window of 301 nt for each position of the reference SARS-CoV-2 genome NC_045512 (https://www.ncbi.nlm.nih.gov/nuccore/NC_045512) and plotted in R (**Fig. S3**). Viral genes that are affected by each probe were identified using the intersect function of bedtools (**Table S3**).

### RT-RPA amplification of viral genomic sequences

Prior to all detection assays a pre-amplification step using RT-RPA was performed in order to amplify the SARS-CoV-2 target sequence. These protocols were initially developed and optimized on mock viral genome fragments and later validated against patient samples. To amplify the target sequences from the synthetic vRNA, we performed RT-RPA ^38^ (protocol summarized in **Fig. S3**). In short, RT-RPA primers were designed to amplify 30 nt gRNA spacer regions flanked by 30 nt priming regions from the synthetic vRNA template while also incorporating a T7 promoter sequence into the 5’end of the dsDNA gene fragment with +2 G’s thus increasing transcription efficiency (**Fig. 1B**) ^90^. RT-RPA was performed at 42°C for 45 min by combining M-MuLV-RT (NEB #M0253L) with TwistAmp® Basic (TwistDx #TABAS03KIT). All RT-RPA primer sequences can be found in **Table S1**.

### Fluorescence-based detection of SARS-CoV-2

For fluorescence-based detection, we developed a simple *in vitro* transcription-coupled cleavage assay with a fluorescence readout using 6-Carboxyfluorescein (6-FAM) as our fluorescent marker. To facilitate fluorescence detection, we developed a 6 nt poly-U probe conjugated to a 5’-6-FAM and a 3’-IABlkFQ (FRU, **Table S5**) and custom ordered from IDT. In total volumes of 15*μ*L, the following reaction mix was prepared; 5.62*μ*L water, 0.4*μ*L HEPES, pH 7.2 (1M), 0.18*μ*L MgCl_2_ (1M), 3.2*μ*L rNTPs (25mM each), 2*μ*L CasRx (55.4 ng/*μ*L), 1*μ*L RNase inhibitor (40U/*μ*L), 0.6*μ*L T7 Polymerase (50U/*μ*L), 1 *μ*L gRNA (10 ng/*μ*L), and 1*μ*L FRU probe (2uM). This was followed by the addition of 5*μ*L (50% amplification vol) of the amplified target RNA from the RT-RPA pre-amplification mix (described above) or no-template control, which initiates the reaction following incubation at 37°C for 90 min. Experiments were immediately run on a LightCycler® 96 (Roche #05815916001) at 37°C under 5 sec acquisition followed by 5 sec incubation for the first 15 min, followed by 5 sec acquisition and 55 sec incubation for up to 75 min. Fluorescence readouts were analyzed over-time by normalization to templateless controls at each respective time point or through background subtracted fluorescence by subtracting the initial fluorescence value from the final value.

### Lateral Flow-based detection of SARS-CoV-2

For lateral flow-based detection we modified the HybriDetect® system to detect the presence of SARS-CoV-2 sequences using SENSR ^38^. In brief, we designed a ssRNA probe composed of a 6 nt poly-U probe conjugated on opposite ends with a 5’-6-FAM and a 3’-biotin which was custom ordered from IDT (LFRU, **Table S5**). Following incubation of 5.22*μ*L water, 0.4*μ*L HEPES, pH 7.2 (1M), 0.18*μ*L MgCl_2_ (1M), 2*μ*L CasRx (55.4ng/*μ*L), 1*μ*L gRNA (10 ng/*μ*L), 5*μ*L RT-RPA reaction mix, 1*μ*L T7 polymerase (50 U/mL), 3.2*μ*L rNTPs (25mM each), 1*μ*L LFRU probe (20 uM), at 37°C for 60 min. 80*μ*L of HybriDetect Assay buffer was added to each reaction and mixed thoroughly. Next, the lateral flow dipstick was placed into the reaction and allowed to flow upwards by capillary action for a maximum of 2 min. The presence or absence of upper or lower bands was analyzed to detect evidence of SARS-CoV-2 by collateral cleavage. The presence of a solitary upper band or both an upper and lower band indicates a positive result, a solitary lower band with a faint upper band was interpreted as a negative result.

### Patient samples ethics statement

Human samples from patients were collected under University of California San Diego’s Human Research Protection Program protocol number 200470 for negatives (PI: Lauge Farnaes), and under a waiver of consent from clinical samples from San Diego County for positives (PI: Kristian Andersen), as part of the SEARCH Alliance activities. Samples were de-identified as required by these protocols prior to testing and analysis under University of California San Diego Biological Use Authorization protocols R1806 and 2401.

### RNA extraction and processing of patient samples

Patient nasopharyngeal samples were collected and RNA was extracted using Omega Bio-Tek Mag-Bind Viral DNA/RNA 96 Kit (Omega Cat. No. M6246-03), following the manufacturer’s protocol for KingFisher Flex platform.

### RT-qPCR validation of SARS-CoV-2 infection in patient samples

Patient samples were determined to be SARS-CoV-2 positive or negative TaqPath™COVID-19 Combo Kit RT-qPCR assay as described in (https://www.fda.gov/media/136112/download), and reducing the RT-qPCR reaction volumes to 3µl and diluting the MS2 phage control to improve the limit of detection of the assay. The presence of SARS-CoV-2 viral RNA was analyzed using primers targeting the N, S, and Orf1ab genes with an MS2 control. All RT-qPCR assays were run using TaqPath™ 1-Step RT-qPCR Master Mix (ThermoFisher #A15299) and thermocycling conditions were run following the CDC recommended protocol (https://www.fda.gov/media/136112/download). Fluorescence data were acquired on a QuantStudio 5 qPCR machine (Applied Biosystems).

### SENSR detection of patient samples

To detect the presence of SARS-CoV-2 in patient samples using SENSR, we tested this system against RT-qPCR validated samples. We obtained SARS-CoV-2 positive (N=21) and negative (N=21) samples and ran fluorescence analysis of these samples in triplicate. Samples were subject to preamplification using RT-RPA and incubated in an IVT-coupled cleavage reaction, as previously described. Data for analysis were acquired on LightCycler® 96 (Roche #05815916001) following the protocol previously described. We processed the data by generating background subtracted fluorescence data for each replicate by subtracting the final (90 min) fluorescence value from the initial (0 min) fluorescence value. Noise was set as the average of the three no-template control (NTC) background subtracted values. S/N was then calculated by dividing the background subtracted value for each replicate by the noise. The S/N for each sample was then determined by taking the average of the three independent S/N ratios in the triplicates. Samples were determined to be positive if S/N > 2 and negative if S/N < 2. Lateral flow analysis was run on samples that were determined as positives from the SENSR fluorescence analysis. The samples were assayed and analyzed following the previously described lateral flow methods and images were taken using a smartphone. Positives and negatives were determined in comparison to the NTC samples and using a positive control (synthetic template) as a standard.

## SUPPLEMENTARY FIGURES

**Supplementary Figure 1.**
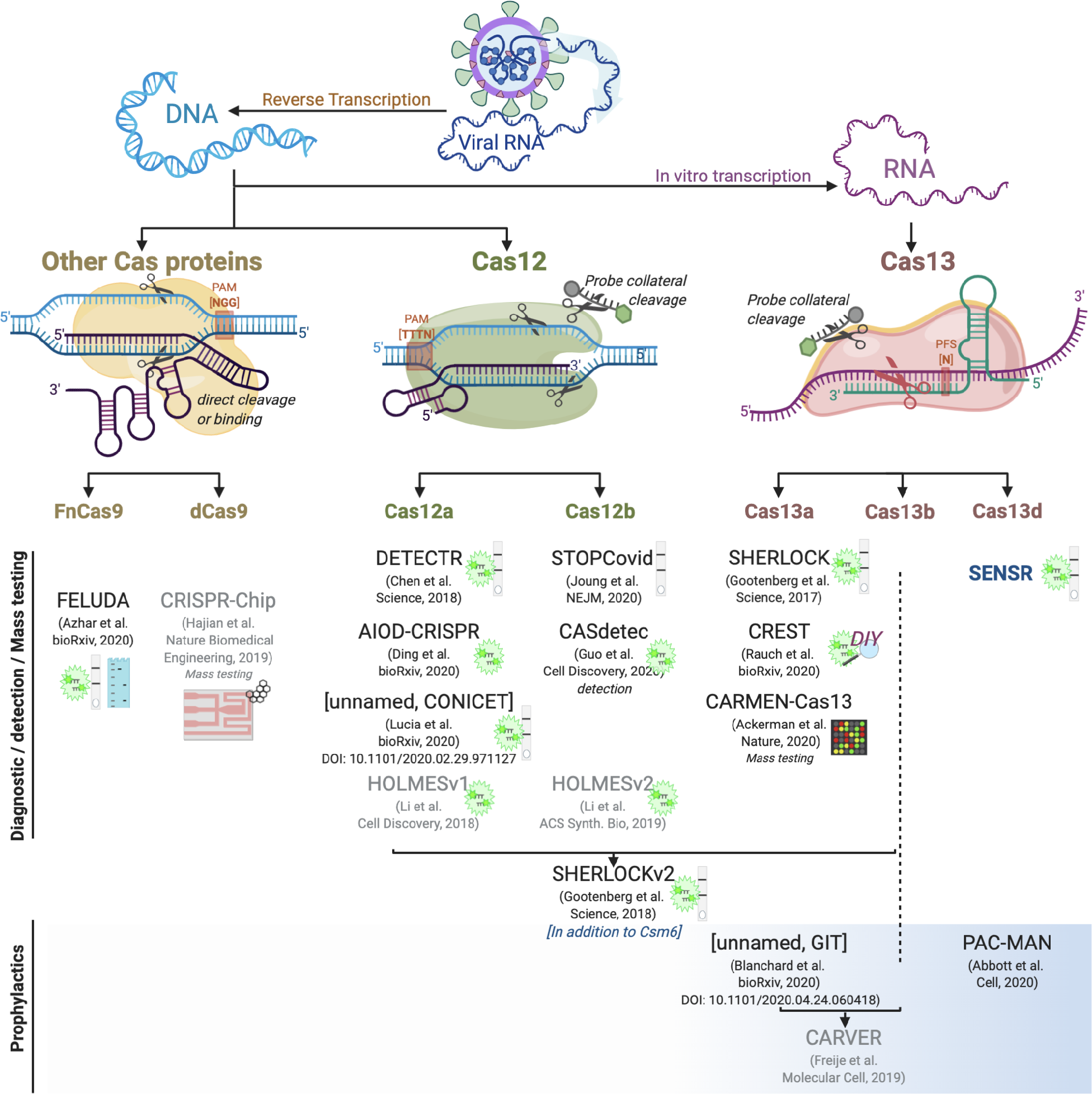
Summary of current CRISPR-based anti-Covid technologies organized by Cas enzyme used and by role as diagnostic or detection tool, or as a putative prophylactic. Those technologies shown in black font have been demonstrated to have explicit activity against SARS-CoV-2, while those technologies shown in grey font have been publicly discussed or proposed to be technological candidates for SARS-CoV-2 detection or prophylaxis, however have not yet been fully demonstrated/optimized for said purpose as of the publication of this manuscript. Some technologies have not yet been given a formal name by their authors, and are therefore denoted as ‘unnamed’followed by the acronym for the primary institutional affiliation behind the work. To better identify and distinguish these technologies, the DOI has been provided. **[Diagnostics/detection systems/Mass testing]** For all technologies viral RNA is extracted, reverse transcribed into cDNA, followed by template amplification by either PCR, RPA or LAMP, then input into subsequent reactions with the exception of CRISPR-Chip which does not require an amplification step. Cas12-based enzymes, as well as many other Cas proteins (including Cas9) recognize DNA species, while Cas13-based enzymes recognize ssRNA, and all can be used to detect evidence of specific sequences by fluorescence or lateral flow. The detection method for each technology is noted with the presence of an icon adjacent. The majority of the technologies summarized here use fluorescence or lateral flow or both. The lime glow icon indicates readout by fluorescence, while the grey bar indicates read-out by lateral flow. Some of the other technologies can be read out by different detection methods. Notably because FELUDA relies on direct sequence cleavage, and not the collateral cleavage property shared amongst the Cas12- and Cas13-based technologies, it can also be analyzed by gel electrophoresis as evidence of distinct band cleavage (aqua gel icon). Also, CRISPR-Chip has been discussed as a SARS-CoV-2 mass detection candidate, though these findings have not yet been made publicly available. Read-out by this technology is achieved via a graphene-based transistor (icon is red, grey with graphene structure adjacent). Other different technologies include CREST, which achieves detection via fluorescence using a distinctly do-it-yourself (DIY) BioHacking approach, by utilizing equipment easily procured and operated by novice scientists, as well as CARMEN-Cas13 which detects evidence of SARS-CoV-2 sequences by microarray. The technology outlined here is an addition to current SHERLOCK detection utilizing Cas13d (CasRx). The technical details of most technologies is summarized in Table S1.

**Supplementary Figure 2.**
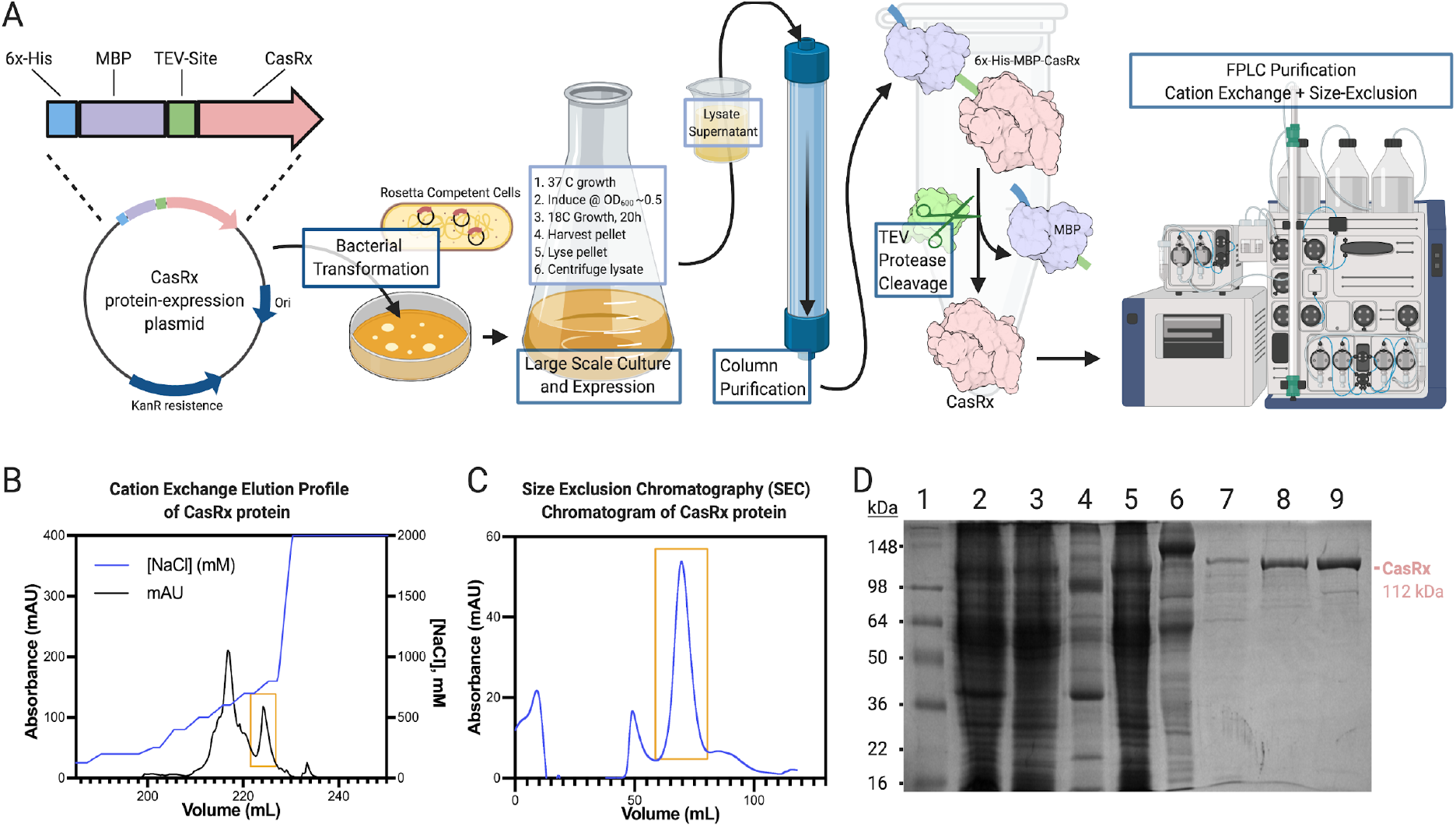
CasRx protein purification workflow and quality control. **[A]** CasRx protein was produced and purified essentially as described in (Konermann et al. 2018). A CasRx protein-expression plasmid was generated with CasRx (pink) downstream of a Maltose-binding protein (MBP, purple) domain with an N-terminal 6xHis tag (blue), connected by a TEV protease cleavage sequence (green). Origin of replication and KanR cassette shown in navy. Transformation of the plasmid into Rosetta2™ (DE3) Competent *E. coli* was followed by large scale culture growth. Cell lysate supernatant containing 6xHis-MBP-CasRx soluble hybrid protein was run on a Ni-NTA column for purification by affinity chromatography. CasRx protein was then released from the 6xHis-MBP moiety by TEV protease cleavage during O/N dialysis, and was further purified by cation exchange and size exclusion using Fast Protein Liquid Chromatography (FPLC). **[B]** The cation exchange elution profile of CasRx with the concentration of NaCl (mM) shown in blue. Peak containing CasRx recombinant protein (boxed in gold) elutes at ∼700 mM NaCl. **[C]** The SEC elution profile of CasRx recombinant protein following Size Exclusion Chromatography (SEC) (blue). Peak containing CasRx recombinant protein highlighted boxed in gold. **[D]** A 10% SDS-PAGE gel showing protein species present at different stages of the purification protocol. Lane 1 is SeeBlue™Plus2 Pre-stained Protein Standard with the predicted final CasRx protein size marked at right in pink at approximately 112 kDa. Lane 2 is a resuspended cell pellet. Lane 3 is cell lysate supernatant. Lane 4 is cell lysate pellet. Lane 5 is Ni-NTA flow through. Lane 6 is Ni-NTA Elution. Lane 7 is the sample post O/N Dialysis. Lane 8 is the sample after IEC, and Lane 9 is the concentrated Final Sample post-SEC.

**Supplementary Figure 3.**
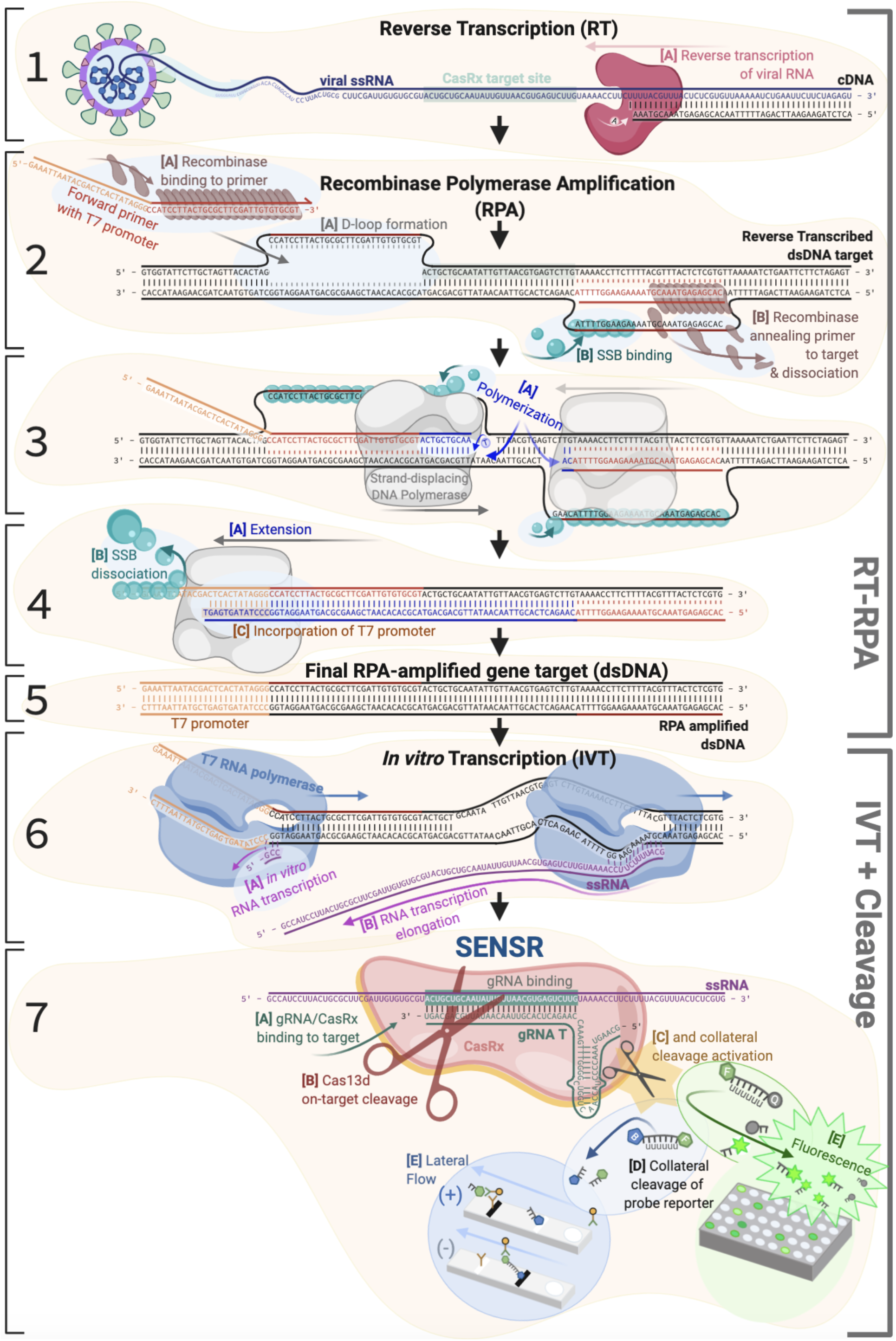
Molecular scale overview of CasRx detection protocol workflow. Bracketed into the two base component reactions, RT-RPA or IVT + cleavage. [**Panel 1A**] Viral ssRNA is extracted and reverse transcribed into cDNA. A fragment of the viral E-gene ssRNA (navy), with the final CasRx target site highlighted (mint). Reverse transcriptase (raspberry) actively reverse transcribes the viral template into cDNA. [**Panel 2A**] In the simultaneous RPA reaction, recombinase (brown) binds to the primer (red and orange) with D-loops formed at binding sites. [**2B**] Recombinase helps anneal the primer to the target site, and single strand binding protein (SSB, teal) begins annealing to ssDNA to stabilize the strand and reaction. [**Panel 3A**] Strand-displacing DNA polymerase (grey) amplifies the target DNA, with continued binding of SSB to ssDNA for stabilization. [**Panel 4A**] DNA amplicon extension is completed, [**4B**] while polymerase simultaneously dislodges SSB, and [**4C**] the T7 promoter region (orange) is added to the amplicon via primer extension. [**Panel 5**] The final product of the RT-RPA reaction is a small amplified fragment of target DNA encompassing the CasRx target site, with a T7 promoter added for subsequent IVT. [**Panel 6A**] T7 RNA polymerase-based *in vitro* transcription (blue) occurs, initiated from the T7 promoter (orange) in order to generate ssRNA (magenta) required as the activation substrate of CasRx. [**6B**] Elongation of the ssRNA template takes place. **[Panel 7**] CasRx detection of on-target sequence, activation of collateral cleavage activity, and detection. [**7A**] The CasRx/gRNA complex (pink and green) recognize, [**7B**] bind to, and cleave (red scissors) the ssRNA viral target template (magenta). [**7C**] This action activates the collateral cleavage property of CasRx (gold, black scissors). [**7D**] Following activation, collateral cleavage of an included small ssRNA probe can be analyzed via [**7E**] fluorescence or lateral flow assay.

**Supplementary Figure 4.**
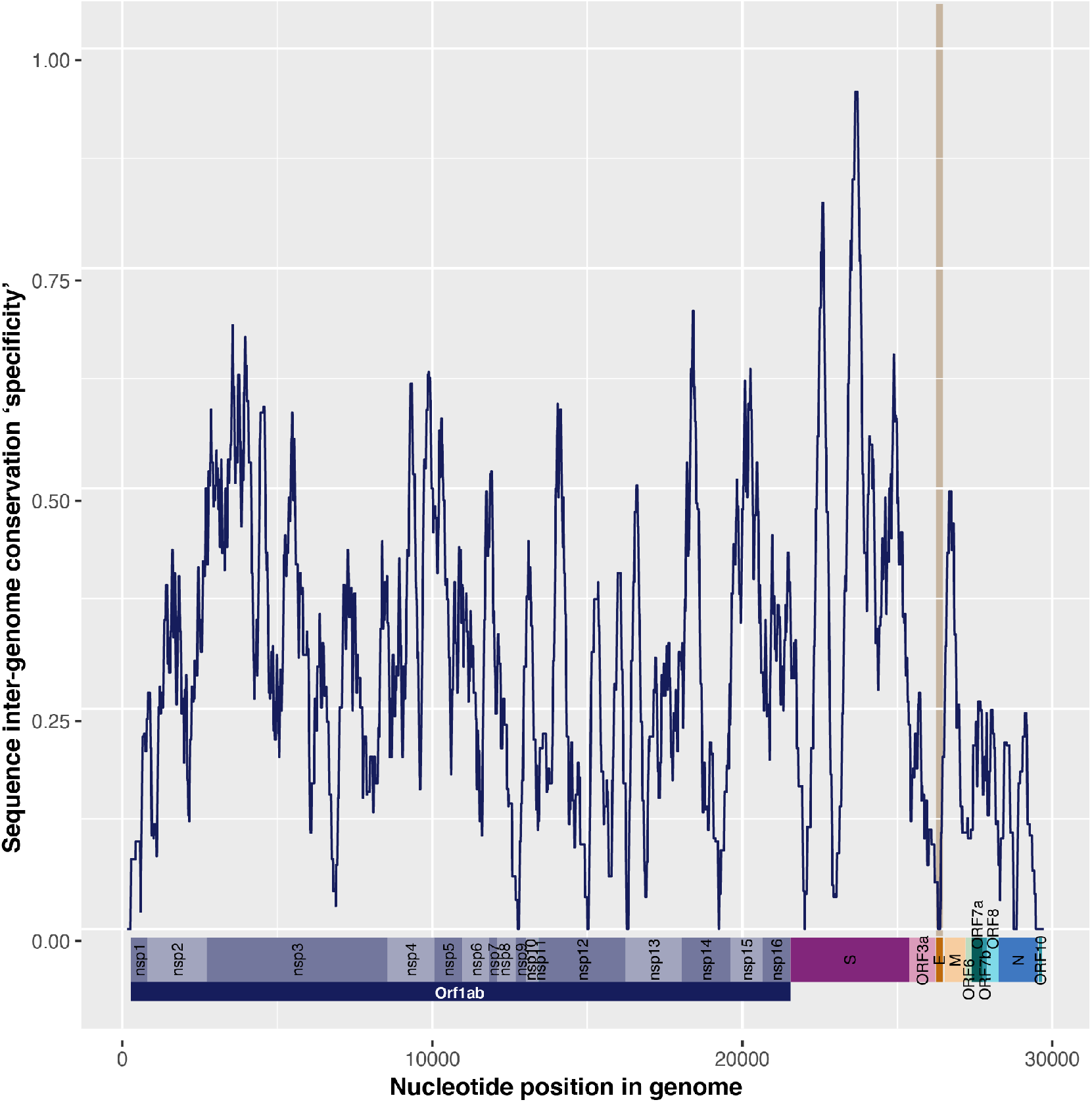
Depiction of unique gRNA target sequences across SAR-CoV-2 genome. Spread of unique and specific 30 nt putative gRNA target sequences (Table S3-S5) displayed across the SARS-CoV-2 genome, smoothened over a 301 nt window. Higher specificity score indicates higher density of unique and conserved targets. Representative genome map shown at bottom with ORFs and genes annotated. E-gene ORF highlighted in orange with beige vertical bar highlighting the zero specificity score of the gene.

**Supplementary Figure 5.**
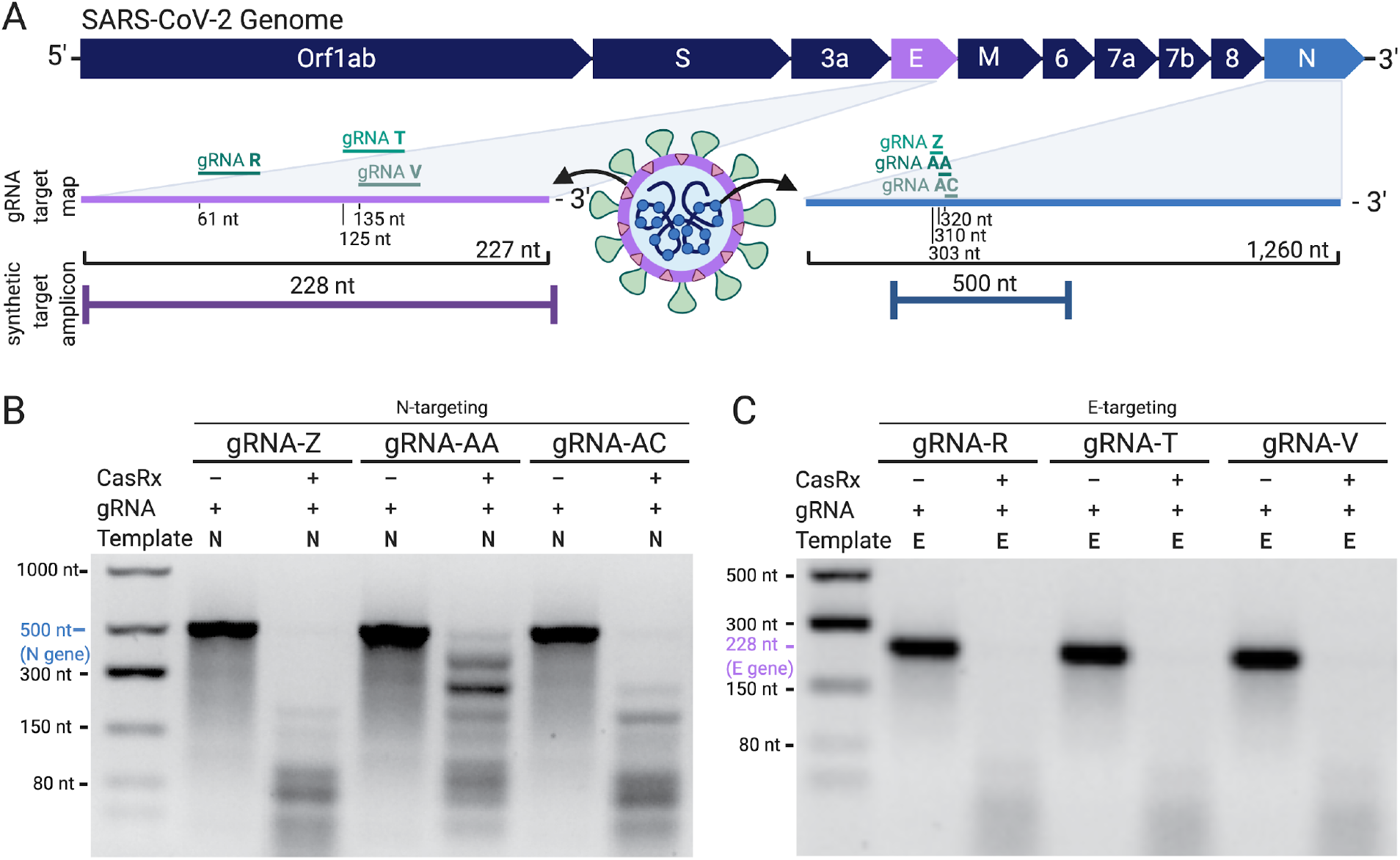
Preliminary *in vitro* cleavage assays for guides tested throughout this work. **[A]** Summary map of gRNA target location within the SARS-CoV-2 genome. gRNA-R, -T, -V target the Envelope (E) gene, and gRNA-Z, -AA, -AC target the Nucleocapsid (N) gene. **[B]** *In vitro* on-target cleavage assays for the six guides developed in the first cohort (R, T, V). Equimolar amounts of N and E synthetic RNA fragments (500 nt and 228 nt respectively) were incubated at room temperature (RT) in the presence or absence of their on-target gRNA, named above. The on-target gene is denoted with (E) or (N) for each gRNA, and is shown mapped in **[A]**. The presence of the gRNA initiates cleavage of both the on-target template in addition to collateral cleavage of the off-target template. **[C]** On-target *in vitro* cleavage assays for the conserved and specific N-targeting gRNA-Z, -AA, -AC. In each reaction the 500 nt N-gene synthetic RNA fragment is provided, and the presence or absence of CasRx initiates on-target cleavage of the template into smaller fragments.

**Supplementary Figure 6.**
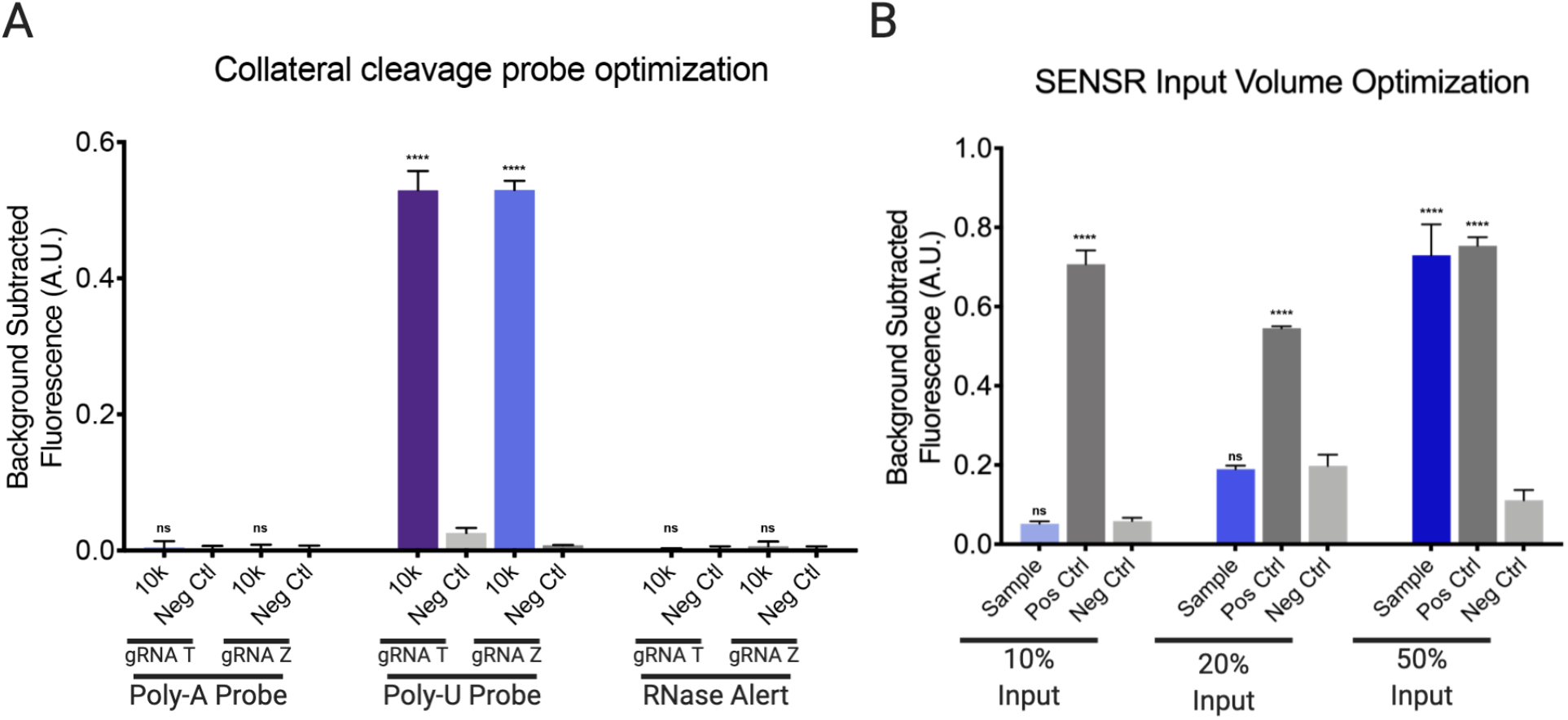
Optimizing SENSR Fluorescence detection assay. **[A]** Determining optimal sequence for collateral cleavage probe. Collateral cleavage of Poly-A probe is tested in the first group, Poly-U is tested in the second, and RNase Alert in the third group. Within each group, gRNA-T and gRNA-Z are incubated in a reaction saturated with 10,000 (10k) target copies, or 0 (Neg Ctrl) as a negative control. Performing a two-tailed t-test of unequal variance comparing each 10k group to the respective Neg Ctrl groups a significant increase in fluorescence was only found for the Poly-U probe 10k groups (gRNA-T 10k: p<0.0001, gRNA-Z 10k: p<0.0001). No significant increase in fluorescence was observed for the 10k Poly-A or 10k RNase Alert groups compared to the respective Neg Ctrl groups for gRNA-T (Poly-A: p=0.5953, RNase Alert: p=0.4294) or gRNA-Z (Poly-A: p=0.7935, RNase Alert: p=0.1510). Performing a one-way ANOVA followed by a Dunnett’s test comparing the 10k groups of the Poly-A and RNase Alert groups to the 10k Poly-U groups a significant difference was found for both gRNA-T (Poly-A: p<0.0001, RNase Alert: p<0.0001) and gRNA-Z (Poly-A: p<0.0001, RNase Alert: p<0.0001). Bars indicate mean ± SD of background subtracted fluorescence from three technical replicates. **[B]** Determining the optimal input volume of RT-RPA sample into SENSR reaction. In all cases the trial sample is equivalent to 100 copies/*μ*L (blue), the positive control is established with 10,000/*μ*L (dark grey), and the negative control is no-template control (pale grey). In the left most grouping the RT-RPA template sample input into the SENSR reaction comprises 10% of the preamplification reaction, in the middle grouping it composes 20% of the preamplification reaction, and in the final grouping it comprises 50% of the preamplification reaction. Performing a one-way ANOVA followed by a Dunnett’s test within each % input group significance was only found for both the sample (p<0.0001) and positive control (p<0.0001) in the 50% group (10%: Sample - p= 0.8925, Pos Ctrl - p<0.0001; 20%: Sample - p=0.8160, Pos Ctrl - p<0.0001). Performing a second one-way ANOVA followed by a Dunnett’s test comparing the 50% sample group to the 10% and 20% sample groups, a significant increase in fluorescence was observed (10%: p<0.0001, 20%: p<0.0001). Bars indicate mean ± SD of background subtracted fluorescence from three technical replicates.

**Supplementary Figure 7.**
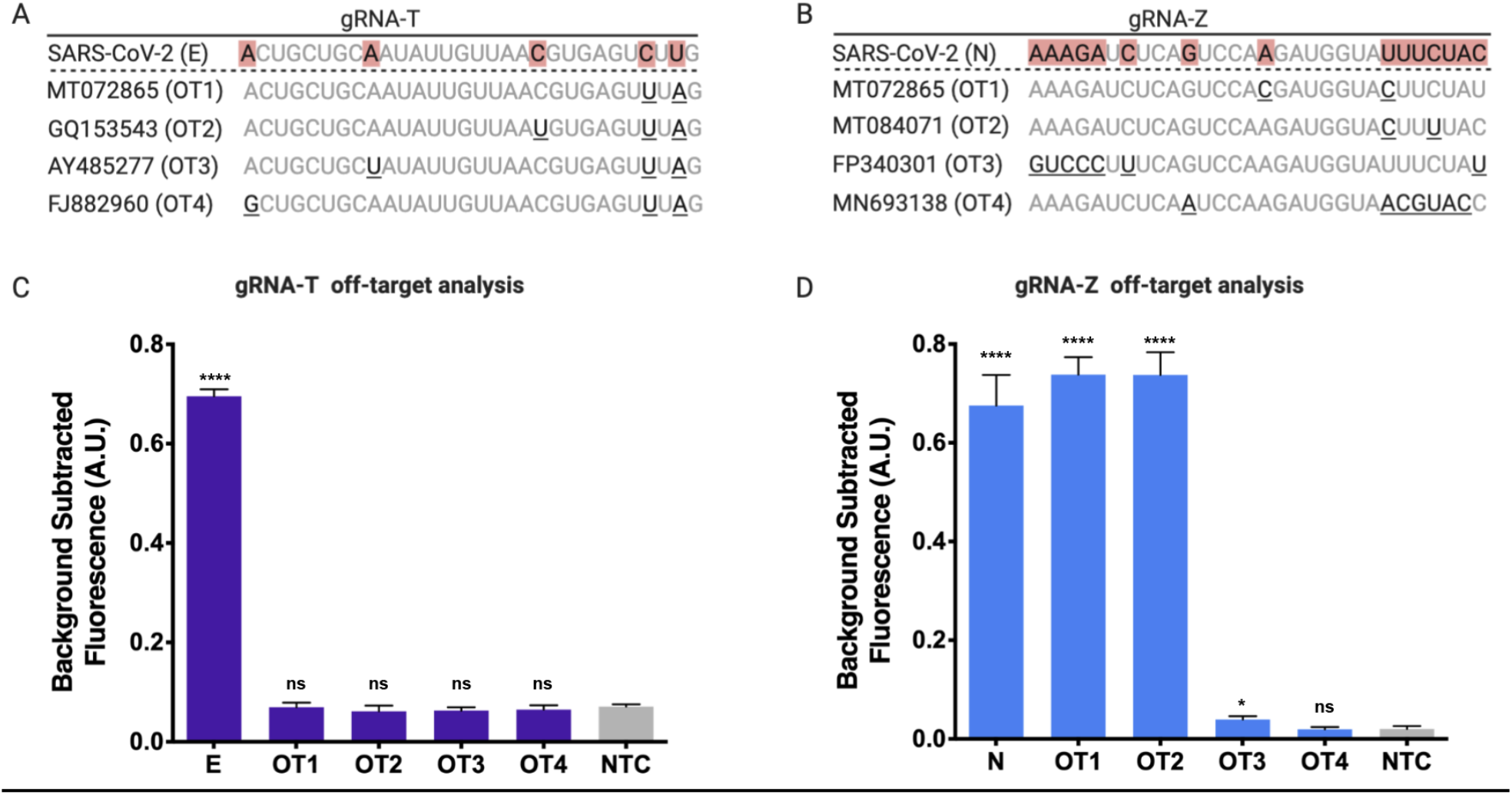
Assessment of gRNA specificity within the SENSR system. Schematic representation of gRNA mismatches for the four most closely related sequences (off-targets, OT1-OT4) identified via BLAST for **[A]** gRNA-T and **[B]** gRNA-Z. **[C]** gRNA-T demonstrated no off-target activity on the 4 closely related sequences. Bars indicate mean ± SD of background subtracted fluorescence from three technical replicates. Performing a one-way ANOVA followed by a Dunnett’s test comparing to the NTC fluorescence signal was found to be significant for the E-gene compared (p<0.0001), however no significant fluorescence signal increase was found for the four off-targets (OT1: p=0.9998, OT2: p=0.5242, OT3: p=0.6633, OT4: p=0.8475). Performing a one-way ANOVA followed by a Dunnett’s test comparing to the E-gene positive control significant fluorescence signal increase was found when targeting the E-gene compared to the four off-targets (OT1, OT2, OT3, OT4: p<0.0001). **[D]** Collateral cleavage was observed for two of the closely related off-targets for gRNA-Z. Bars indicate mean ± SD of background subtracted fluorescence from three technical replicates. Performing a one-way ANOVA followed by a Dunnett’s test comparing to the NTC fluorescence signal increase was found to be significant for the N-gene positive control, OT1, and OT2 (N, OT1, OT2: p<0.0001), however no significant fluorescence signal increase was found for OT3 and OT4 (OT3: p=0.8877, OT4: p>0.9999). Performing a one-way ANOVA followed by a Dunnett’s test comparing to the N-gene positive control, significant fluorescence signal increase was found to when targeting the N-gene compared to OT3 and OT4 (OT3, OT4: p<0.0001), but no significant difference was found targeting the N-gene compared to OT1 and OT2 (OT1: p=0.0786, OT2: p=0.0833).

**Supplementary Figure 8.**
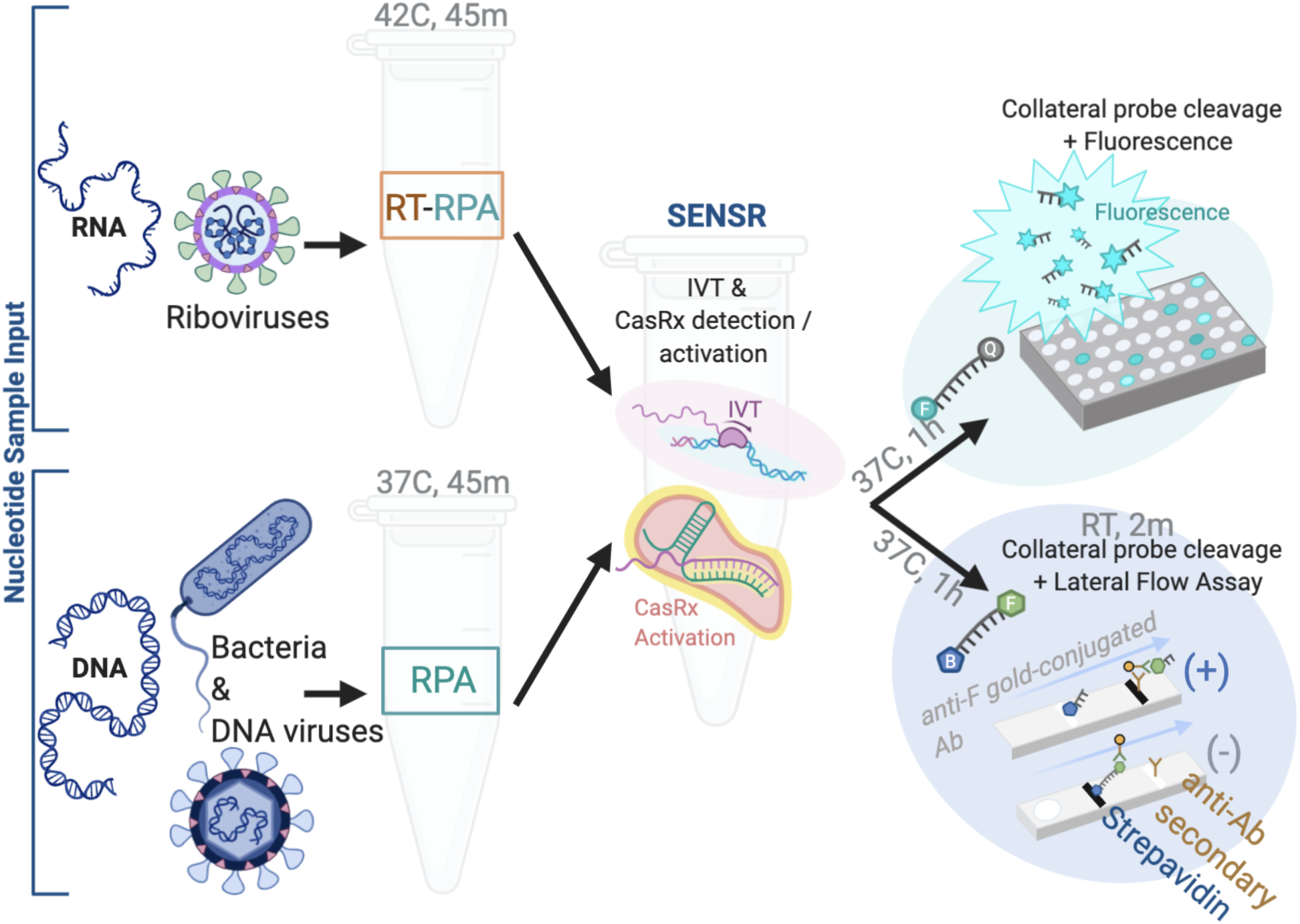
In addition to RNA-based templates (top row), SENSR technology could also be used for DNA-based templates, including, but not limited to, bacterial samples (shown) as well as DNA viruses such as herpes (shown, bottom, navy). DNA-based sample templates can be input directly into the RPA amplification reaction, negating the need for a simultaneous reverse transcription (RT) reaction as is required for RNA-based templates. Like SENSR on RNA-based templates, the reaction occurs in two steps. The first amplification step, RT-RPA or RPA, is followed by the SENSR reaction which is identical regardless of initial input molecular species. This reaction is composed of simultaneous *in vitro* transcription (IVT) as well as CasRx cleavage and activation of collateral cleavage activity. The assay can be read out by fluorescence or lateral flow in the same fashion regardless of DNA or RNA sample input.

## Supplementary Tables

**Table S1**. Summary Table of current, and possible future, CRISPR-based anti-Covid technologies.

**Table S2**. Analysis identifying putative 30nt gRNA target sites conserved across, and specific to the SARS-CoV-2 genome.

**Table S3**. Predicted unique and conserved 30nt CasRx gRNA target sequences to SARS-CoV-2.

**Table S4**. Analysis of Inter-SARS-CoV-2 Conservation (433 genomes) and Pan-coronavirus Specificity (3164 genomes) on the three E-targeting gRNAs (R,T,V).

**Table S5**. List and sequences of reagents generated and used throughout this work. Primers for cloning, gRNA prep, and RT-RPA, as well as gRNA sequences, viral gene templates, plasmid sequences and probes.

**Table S6**. Top four naturally-occurring off-target sequences for gRNA T and gRNA Z.

**Table S7**. Data from RT-qPCR and SENSR fluorescence analysis of patient samples for detection of SARS-CoV-2.

